# Estimating Effect-sizes to Infer if COVID-19 transmission rates were low because of Masks, Heat or High because of Air-conditioners, Tests

**DOI:** 10.1101/2021.01.15.21249896

**Authors:** C. K. Sruthi, Malay Ranjan Biswal, Brijesh Saraswat, Himanshu Joshi, Meher K. Prakash

**Author notes:** contributed equally (C. K. Sruthi, Malay Ranjan Biswal). contributed equally (Brijesh Saraswat, Himanshu Joshi).

## Abstract

How does one interpret the observed increase or decrease in COVID-19 case rates? Did the compliance to the non-pharmaceutical interventions, seasonal changes in the temperature influence the transmission rates or are they purely an artefact of the number of tests? To answer these questions, we estimate the effect-sizes from these different factors on the reproduction ratios (R_t_) from the different states of the USA during March 9 to August 9. Ideally R_t_ should be less than 1 to keep the pandemic under control and our model predicts many of these factors contributed significantly to the R_t_’s: Post-lockdown opening of the restaurants and nightclubs contributed 0.04 (CI 0.04-0.04) and 0.11 (CI. 0.11-0.11) to R_t_. The mask mandates helped reduce R_t_ by 0.28 (CI 0.28-0.29)), whereas the testing rates which may have influenced the number of infections observed, did not influence R_t_ beyond 10,000 daily tests 0.07 (CI -0.57-0.42). In our attempt to understand the role of temperature, the contribution to the R_t_ was found to increase on both sides of 55 F, which we infer as a reflection of the climatization needs. A further analysis using the cooling and heating needs showed contributions of 0.24 (CI 0.18-0.31) and 0.31 (CI 0.28-0.33) respectively. The work thus illustrates a data-driven approach for estimating the effect-sizes on the graded policies, and the possibility of prioritizing the interventions, if necessary by weighing the economic costs and ease of acceptance with them.

## 1. INTRODUCTION

In March-April of 2020, most countries had adopted a partial or complete lockdown in response to the growing COVID-19 pandemic. The lockdowns were mainly motivated by the rapidly rising infections, and a lack of preparedness for the critical healthcare equipment. The added justification came from epidemiological models which projected severe loss of lives for several scenarios of basic reproduction rates (R_0_) characterizing the spread of infections and the case fatality rates [**1,2**]. Subsequent counterfactual models after the lockdowns underscored the reduction in the time varying reproduction rates (R_t_) [**3**] and loss of lives averted by the lockdowns and provided a *post hoc* justification.

Within a month of these partial or full lockdowns, governments began graded re-openings to contain the economic losses. With no vaccines in sight despite intense research efforts [**4**], WHO earlier warned the countries of resurgence if they opened up too soon. Even though vaccines are available now, it will require many months to perform mass-vaccinations. As feared, there were resurgences in many countries. Several states and even cities had autonomously implemented restrictive measures over-riding the decisions at the national or state level, and even entering into legal battles on these issues. These autonomous decisions were based on first-hand information about the clusters of infections stemming for example, from night clubs or because of the understanding that masks prevent the dispersion of the droplets.

In the pandemic situation, the immediate attention of the public or the governments concerned about critical healthcare resources is drawn to the number of daily new infections and cumulative infections. However, these infections are a consequence of the transmission or reproduction rates. While the basic reproduction ratio of COVID-19 is estimated to be around 2.5 to 4, it reduced considerably during and after the lockdowns. Several non-pharmaceutical interventions have been enforced [**5**]. However the factors accounting for this reduction in R_t_ remain elusive, even at the broader level of government policies, testing rates and seasonal effects. While this causal relation is implicitly understood and assumed to be true, predicting how the policies translate to the transmission rates has remained a missing link in data-driven or evidence-based decision making. Data driven or evidence-based decision making by exploiting the data and artificial intelligence are being hailed as the next frontiers in many areas, such as public policy making [**6**] or medicine. However, in the COVID-19 pandemic, artificial intelligence had been used mainly for surveillance and for diagnostics, and not for policy making.

Many epidemiological predictions studied the consequences of different R_t_’s and highlighted the need to keep it in control [**7**]. However, the epidemiology of COVID-19 pandemic had been quite unique [**8**], as it defied the intuitions on infectiousness period, pre-symptomatic and asymptomatic transmission [**9, 10**] of most respiratory infections, including SARS 2003. How the different non-pharmaceutical interventions can reduce the rates to an acceptable level was not clear and each government used a different combination of the different non-pharmaceutical intervention policies. There have been no predictive models translating the policy decisions to the transmission or reproduction rates.

Predicting the rates requires training a mathematical or computational model on sufficiently large and systematized data that contains the information on the policies and their consequences. In the initial days of the pandemic, one could find universalities in the rise of infections across the different countries [**11, 12**]. However, as the policies and the trajectories diverged, this ability to compare or combine data from across the different countries had been lost. A few studies attempted to pool together the learnings by focusing on the reproduction rates which can be compared across different situations. The effectiveness of policy measures from the data pooled from several European countries during February to May [**13**] demonstrated a significant reduction in the reproduction rate due to lockdown. However, many of the interventions in March were implemented in rapid succession and their individual effects could not be decoupled. Other studies reported time-varying reproduction numbers with a detailed state-level tracking in United States [**3**], and correlated them to the mobility patterns [**14**].

In this work, we build a predictive model by pooling together the policy and infection data from the 50 states of United States in the 9 March – 9 August period. Through this period, these states witnessed pre-lockdown and lockdown phases along with a diverse combinations of policy instruments ranging from a graded re-openings of restaurants, bars, to imposition of mask mandates. An implicit assumption is that the different states of the USA are similar in many ways including testing policies and rigor of policy implementation, and that there is an opportunity to learn from other states which experienced same or similar situations. The model which is developed using the data from different states can of course be refined on a continuous basis by also including the standardized data from different countries.

The aim of this work is to introduce a data-driven approach for deciphering the effect sizes of various factors which contribute to the spread or mitigation of COVID-19: firstly, by showing that the transmission rates can be predicted from the knowledge of standardized policies, and then by demonstrating the possibility to decouple the relative contributions of the individual policy instruments.

## 2. RESULTS AND DISCUSSION

### 2.1. Model for estimating the effect-sizes

#### 2.1.1. Standardization for data-driven policies

A data-driven approach will require a standardized set of the different non-pharmaceutical instruments and their measurable consequences. For the latter, we use the transmission rates, which depends on how people interact with each other and is independent of number of current number of infections. Most state governments of USA used various combinations of the policy instruments such as the degree of opening of the retail stores, restaurants, bars, personal care businesses such as barber shops, nightclubs, child care, places of entertainment such as movies, allowed size of gatherings, places of worship, beaches along the oceans or lakes, and outdoor activities. The policy of the states on quarantine restrictions upon inter-state arrivals, and the rigor of mask requirement were also noted and used as independent variables. 14 different policy instruments and the average mobility from Google [**14**] each week were used as the independent variables. These degrees of opening were codified as discussed in the **METHODS** section. For example, the codes we assigned to the restaurant opening policies were: 0 (completely closed), 1 (allowed for a takeaway), 2 (allowed up to 25% seating capacity), 3 (allowed up to 50% of seating capacity), 4 (allowed for more than 50% seating capacity), 5 (pre-lockdown occupancies). A true pre-lockdown situation, with a 100% capacity and no frequent hand washes or use of personal sanitizers, may not be reached in the near future.

The government policies on graded closing or re-openings were gathered from the press releases of the Governors of the 50 states of USA. We discretized the period under study into weeks. For simplicity we follow the standard week numbering convention that identifies the week starting 9^th^ March 2020 as the week 11. If a policy change was implemented anytime during the week, the code for the new policy was uniformly applied throughout the week.

In reality, a restaurant may not be populated, despite a relaxation of the restrictions and additional data will be required to understand these subtle differences between allowance and compliance. However, in this work we assigned the codes to all the independent policy variables based on the limits allowed by the governments. Whether or not the restrictions of gatherings were applicable to the places of worship was not considered. Also, during this period starting from spring break till the summer, schools were closed throughout the United States and hence it was not considered either in our present model, although newer situations and policies may be added in future.

#### 2.1.2. Transmission and Reproduction Rates

Reproduction ratios are standard epidemiological descriptors, which clarify the relative rates of transmission (α_t_) and recovery from the infection (γ). The daily transmission rates were calculated as 1/I_active_. (dI/dt) where I_active_ is the number of active infections, and dI/dt is the number of daily new infections on a given day. The daily new infection data was obtained from Worldometer [**15**]. We work with the assumption that the function is piece-wise continuous [**13**], with the resolution of a week [**12**]. The average transmission rate for the week was calculated, α_t_=<1/I_active_. (dI/dt)>.

The recovery rates (γ) for each state was estimated by following the turn-arounds or the peaks in the number of daily new infection and the results presented are based on these individual state γ estimates. γ_av_ of the different states, 1/γ_av_=15.92 days for all the states corresponds well with the overall 24 days [**16**] and the 15 days for people requiring medium-care [**17**] reported in the literature. It must be noted that the final results did not change even when R_t_ was estimated using the average γ_av_ of the different states, although the results presented in this work use the individual γ’s.

#### 2.1.2. Machine learning Model to Relate the Determining Factors to Reproduction Rates

Our goal in this work is to show that the transmission rates or alternatively the reproduction rates can be predicted from the policy decisions, by learning from same or similar experiences from all states. Considering a median incubation period of around 5-6 days [**18**], the policies adopted in a week are assumed to affect the transmission rates observed in the following week. The weekly policy data from 50 states from 9 March (Week 11) to August 2 (Week 31) were used as independent variables to predict the observed rates in the weeks 12 to 32. We developed an artificial intelligence model based on XGBboost for this purpose (**METHODS**). 14 different policy instruments and the average mobility from Google [**14**] each week were used as the independent variables. The hyperparameters were optimized using the RandomSearch, and calculations performed in Python (**METHODS**).

Seven states never issued a stay-at-home order, although businesses were closed. Others did so at different times between 16 March-31 March, while beginning some recommendations or restrictive measures. Thus, it was not easy to codify the policy data especially until March 31. We thus performed the calculations with two scenarios, one assuming the states were not applying restrictions until they announced the stay (or safe) at home order, and another from the day the graded shutdown recommendations were made in mid-March. Using either of the data sets we obtained similar results. In both cases we trained the model on 80% of the 1071 data points from 51 states and 21 weeks and tested it on the remaining 20%. We obtained comparable results (R^2^_training_ = 0.79, R^2^_test_ = 0.76 for both), indicating that predictions of the rates are robust to some differences in interpretation of policy. The complete set of results are in the **Supplementary Information**. In this work we only discuss the first scenario. The model we developed using the policies adopted by the different USA states demonstrates the predictability of the transmission rates from the knowledge of the various factors including the policies and weather conditions. Further, we believe the approach which predicts the transmission rates to be observed in the following week also implicitly suggests how quickly one can expect to see the results of major policy changes.

#### 2.1.4. Decoupling the Contributions from the Different Policy Instruments

Since various combinations of these different policy instruments were used simultaneously it had not been easy to decouple their relative effect sizes. We performed a post-processing of the model predictions using an interpretable artificial intelligence framework known as the SHapley Adaptive exPlanation (SHAP) [**19**]. SHAP analysis of the R_t_ predictions provides the contribution of each factor, from state in each week of calculation. The collected SHAP values for each degree of policy measure are then used for performing paired t-tests among the different factors.

### 2.2. Effect sizes

#### 2.2.1. Graded openings of Dine and Drink businesses

The Governors of various states decreed the permission to open the restaurants in various degrees take-away only (code 1), 25% of dining outdoors (code 2), 50 % of the restaurant capacity (code 3), unrestricted (code 4), pre-lockdown level (code 5). The contributions of this factor to the R_t_ in the various states, and across the weeks is shown in Figure 1A. The effect size between having the restaurants completely closed (code 0) and allowing an unrestricted opening (code 4) is estimated to be 0.04 (CI 0.04-0.04) (Table 1). There was no significant difference if the restaurant was allowed at 25% of its capacity. We believe that practically a complete pre-lockdown scenario may not arise in the near future and hence was not used in the effect-size estimations. Similar analyses for the bars (Figure 1B) and nightclubs (Figure 1C) allowed us to estimate the effect sizes to be 0.11 (CI 0.11-0.11) and 0.16 (CI 0.15-0.16) respectively.

**Table 1.** Effect sizes of different variables used in the model. The effect size on the time varying reproduction rates (R_t_) between the lockdown and the most relaxed post-lockdown scenario are shown for all the variables used in the analysis estimated using paired t-test. *The variables have a decreasing effect on R_t_. ^†^The comparisons of the mobility are between the lowest mobility during lockdown, and the intermediate mobility which had the highest impact after lockdown. ^§^ Daily testing rate comparisons are between the test rates of 10,000 per day versus 30,000 per day.

| Index | Variable | Effect Size on $R_t$ | Index | Variable | Effect Size on $R_t$ |
| --- | --- | --- | --- | --- | --- |
| Non-pharmaceutical intervention policy |  |  | Non-pharmaceutical intervention policy |  |  |
| 1 | Mask mandate* | 0.28 (CI 0.28-0.29) | 12 | Outdoor sports permits | 0.01 (CI 0.02-0.05) |
| 2 | Restaurant opening | 0.04 (CI 0.04-0.04) | 13 | Gatherings with social distance | 0.04 (CI 0.03-0.05) |
| 3 | Bar opening | 0.16 (CI 0.15-0.16) | 14 | Entertainment & Theaters | 0.06 (CI 0.06-0.06) |
| 4 | Nightclubs opening | 0.11 (CI 0.11-0.11) | 15 | Worship | 0.55 (CI 0.54-0.56) |
| 5 | Mobility† | 0.38 (CI 0.31-0.45) |  |  |  |
| 6 | Travel Restrictions* | 0.05 (CI 0.04-0.05) | Testing rate policy |  |  |
| 7 | Beach Access | 0.07 (CI 0.07-0.07) | 16 | Daily Testing Rate§ | 0.07 (CI -0.57-0.42) |
| 8 | Retail Opening | 0.06 (CI 0.06-0.07) |  |  |  |
| 9 | Childcare & summer camps | 0.01 (CI 0.00-0.01) | Temperature factor |  |  |
| 10 | Barbers, Salon, Spa opening | 0.11 (CI 0.11-0.11) | 17 | Cooling needs (CDD55) | 0.24 (CI 0.18-0.31) |
| 11 | Gym & Athletics opening | 0.06 (CI 0.05-0.06) | 18 | Heating needs (HDD55) | 0.31 (CI 0.28-0.33) |

**Figure 1.**
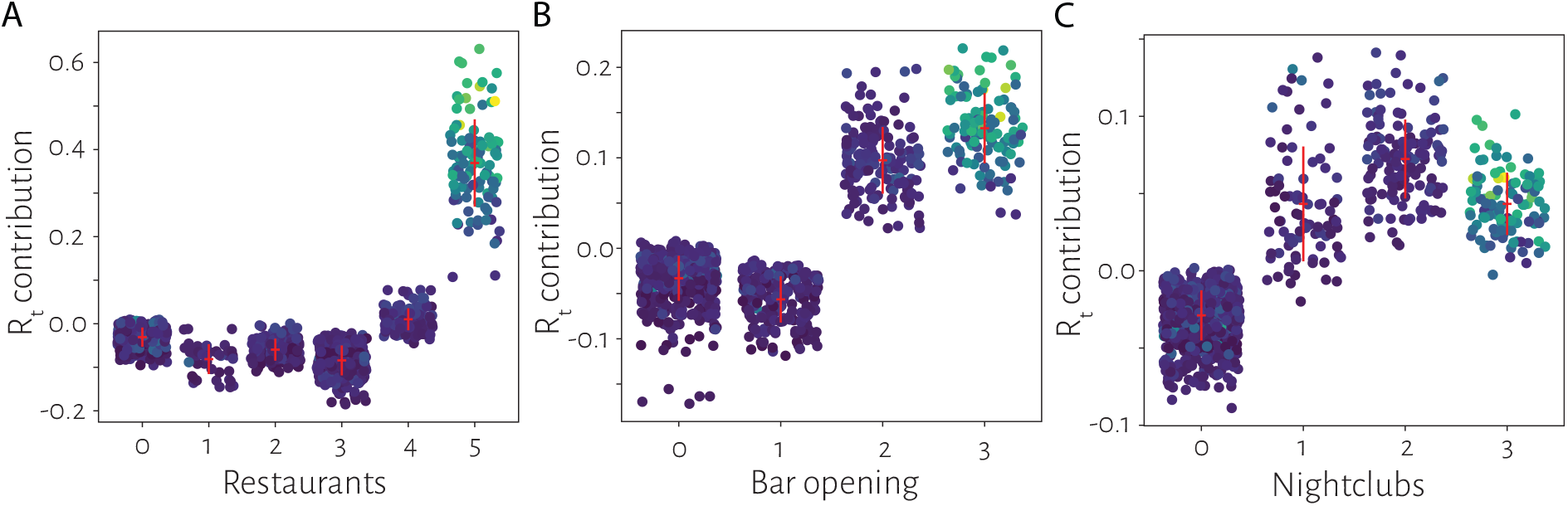
Effect sizes from dine and drink businesses. The contributions of the graded openings of A. Restaurants B. Bars C. Nightclubs are shown. Each point indicates the contribution of the specific factor to the predictions of R_t_, indicated by the color, in the state and week combination. The red markers indicate the mean and standard deviations for each group. To estimate the effect sizes, the pre-lockdown level which is represented by the highest value in these categories was not included.

#### 2.2.2. Mobility related factors

The effect sizes from the mobility estimated using the Google mobility data suggests a saturation in the contribution at 0.38 (CI 0.31-0.45) (Figure 2A). This is plausible because a moderate amount of mobility after a complete lockdown period had its effects. However, beyond a certain degree the mobility was not a direct contributor. The effect sizes of imposing a quarantine upon arrival or allowing access to beaches was estimated to be 0.05 (CI 0.04-0.05) and 0.07 (CI. 0.07-0.07) respectively.

**Figure 2.**
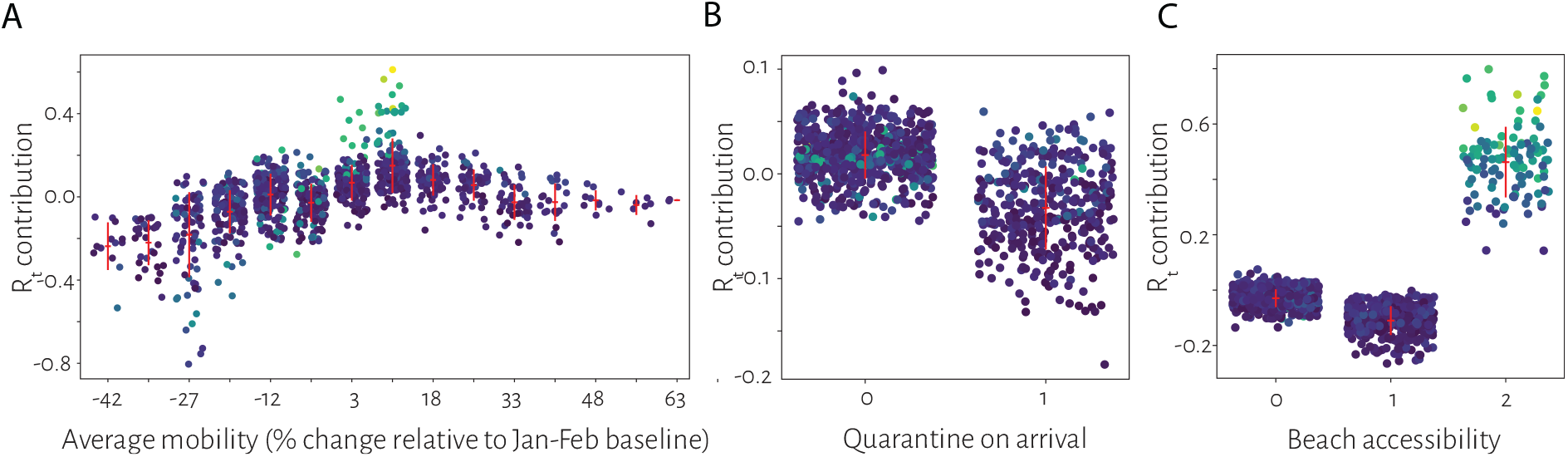
Effect sizes from mobility. Contribution from the different aspects of mobility A. relative movement as measured by Google mobility data B. Enforcement of quarantine upon arrival from an interstate travel C. Accessibility of beaches. The mobility played a role when the lockdown was released

#### 2.2.3. Enforcement of mask-mandates and testing rates

The efficacy of the use of masks is typically justified from laboratory experiments. Here by comparing the weeks with and without mask mandates (Figure 3A), we estimate the effect size respectively to be 0.28 (CI. 0.28-0.29) and 0.07 (CI -0.57 to 0.42) for testing rates 10,000 vs. 30,000. The latter suggests that the lower rates of testing may not mask the reproduction rates as long as they exceed a threshold.

**Figure 3.**
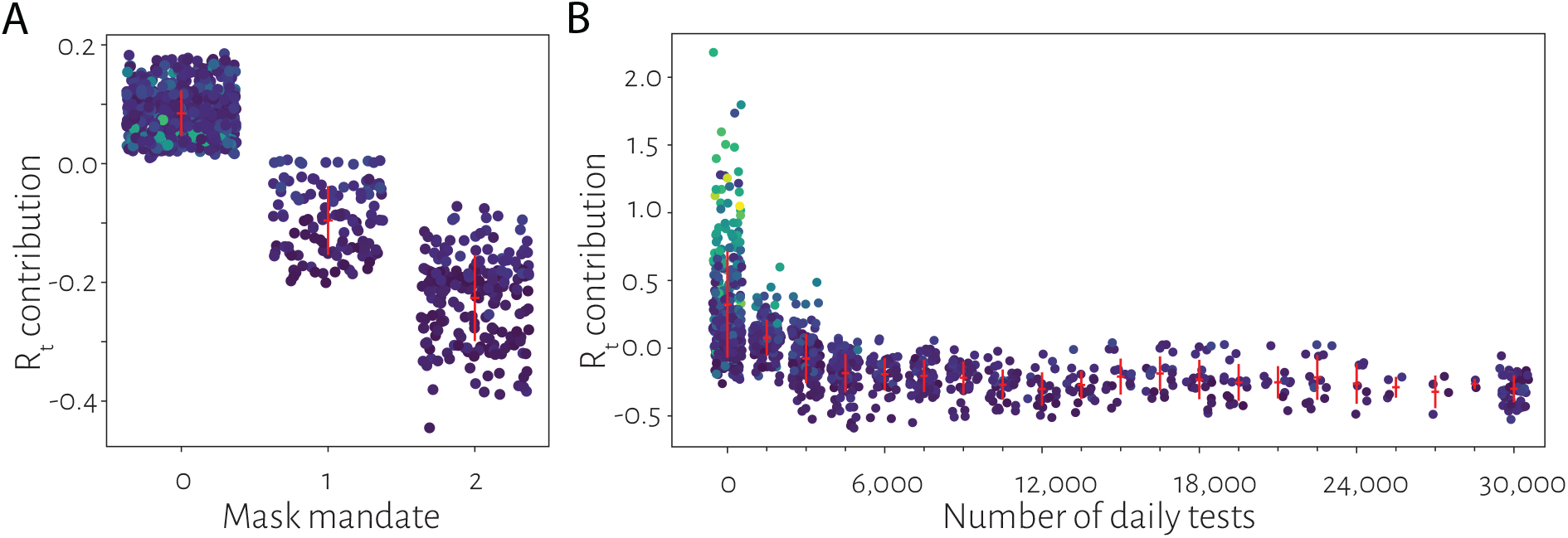
Mask Policies and Testing rates. The contribution of the A. mask mandate and B. number of daily tests to the transmission rates is shown. To estimate the effect sizes, no mask recommendation (Code 0) was compared with a mask mandate either at the state level or in many of the most populated counties (Code 2) was used. The effect size of testing rate was obtained by comparing 10,000 daily tests with 30,000 daily tests.

#### 2.3.4. Effect of temperature

To understand the seasonal effects, temperature and humidity were used as an additional variable in the model. Our analysis resulted in a non-monotonous (Supplementary Figure 1) dependence on temperature with a minimum at around 55F. A minimum at 55F is typically observed in the state level residential energy consumption. We hypothesize that climatization needs of using airconditioners above 55F and heating below 55F, rather than the climate itself may be the major contributing factors. Following this hypothesis, we repeated the calculations by replacing the temperature with two variables – heating degree days relative to 55F (HDD55), and cooling degree days 55F (CDD55) which are the standard ways of measuring energy requirements for heating and cooling. As the results in Figure 4 show, the effect of the need for cooling and heating capture 0.24 (CI 0.18-0.31) and 0.31 (CI 0.28-0.33) contributions to the R_t_.

**Figure 4.**
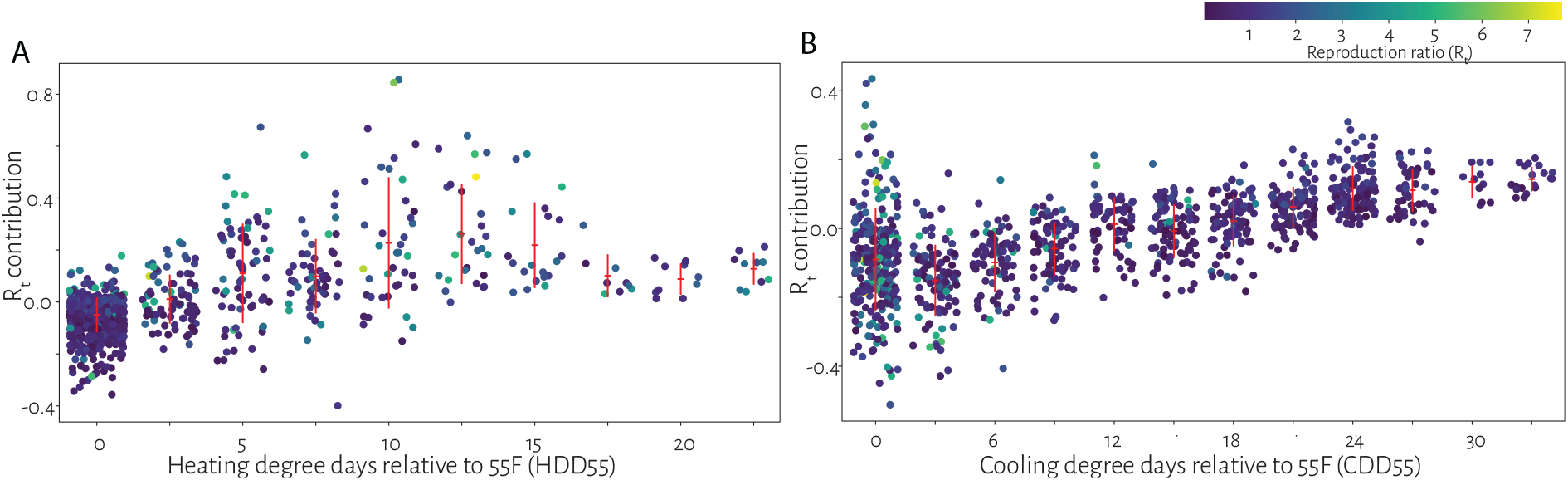
Contribution of heating or cooling needs to the reproduction ratio. The contribution of the use of air-cooling to transmission rates. Cooling degree days with 55F (CDD 55F) as a reference temperature is indicative of the energy needs for cooling, and in this work it is believed to be a surrogate for the use of air conditioners.

#### 2.3.5. Contributions from other factors

The contributions to the various factors such as the opening of the barber shops, childcare, retail stores and gyms (Supplementary Figure 2) allowing of outdoor sports and gatherings (Supplementary Figure 3) or opening of places of entertainment and of worship (Supplementary Figure 4) were also estimated (Table 1).

##### Integrating More Data from Newer Scenarios

The policies adopted by the states of USA had been quite different. Although most of them responded to the national emergency with lockdowns, the reopening strategies were quite different, at times appearing random with no clear rational basis. However, purely from the modelling perspective the ensemble of policies the governments adopted help in training better models. Fine-graining the data from different counties in a state, and following their individual policies and rates will serve the dual purpose of removing regional heterogeneities in rate estimation and increasing the data available for the analysis. The estimation of the effect sizes, as we did here, allows one to weigh the risks of increasing the R_t_ with the economic losses or general public inconvenience, to make informed decisions.

## Conclusions

In summary, we show that a predictive model could be built by integrating the data from different locations, despite the differences in the policy landscape. Standardizing the data from more geographies, using refined tools of AI, one will be able to improve the predictions and interpretations about the role of the policy instruments on the COVID-19 transmission rates, providing guidelines for a data-driven policy-making.

## METHODS

### Policy data curation

The information about the policy data was curated from the official press releases of the Governors of the states and by cross-checking them with the media reports. The following codes were employed:

- *BarbersSalonSpa*: Everything closed (0), Beauty salons, Barbers opened (1), nail spas, tanning salons, tattoo parlors also allowed to operate (2), Prepandemic level (3)
- *Bars*: Bars are places which primarily sell alcohol and not food. Closed (0), Opened with 25% capacity (1), Opened with 50% capacity (2), Pre-lockdown level (3)
- *Beaches*: Beaches on oceans such as in California or Lakes in states without ocean but still attracting tourists were considered. Beaches closed (0), open with severe restrictions (1), open with lesser restrictions (2), Pre-lockdown level (3)
- *Childcare*: Closed (0), Open (1), Summer camps allowed as well (2), Pre-lockdown level (3)
- *CDD55 and HDD55*. The metrics of the amount of cooling or heating required was obtained from http://www.degreedays.com
- *Gathering size*: The allowed gathering sizes were used - 0, 3, 5, 9, 10, 25, 50, 100, 200, 250 (when it was mentioned as 250 or unrestricted)
- *Mask*: No mask requirement (0), Mask usage is recommended but not mandatory or mandatory in some cities (1), Mask usage is mandatory (2)
- *Mobility*: Mobility data relative obtained from Google COVID-19 Community Mobility [**Google**] Reports, and was used as an average of all the categories that were given
- *Nightclubs*: Closed (0), Opened with 25% capacity (1), Opened with 50% capacity (2), Pre-lockdown level (3)
- *Outdoor sports*: Stay at home with no outdoor activities (0), Parks, tennis courts, dog parks, hikes allowed and also Pre-lockdown level (1)
- *Restaurants*: Closed (0), Takeaway (1), fractional capacity of up to 25% (2), fractional capacity of up to 50% (3), Unrestricted (4), Pre-lockdown level (5)
- *Retail*: All non-essentials closed (0), partial opening (1), extended opening including books, florists, etc. or auto showrooms (2), Pre-lockdown level (3)
- *Schools*. Hybrid model.
- *Travel restrictions*: No quarantine restrictions upon interstate arrival and also Pre-lockdown level (0), Inter-state arrivals from some or all states are asked to quarantine or test upon arrival (1)
- *Worship*: Places of worship such as Churches closed (0), open with restrictions (1), Pre-lockdown level (2)
- *Temperature and Humidty*. The temperature and humidity data was obtained from https://www.weatherunderground.com
- There were other state specific parameters such as opening of movie shootings in California, high spread of infections in the meat industry in Nebraska. All these state specific parameters were not considered. The number of tests conducted could be another variable. However, we did not use it in the present model.

### AI model

We used the *XGBoost* library in Python for developing an AI model to predict the rate. 80% of the complete data set consisting of 1071 data points, was used as the training set and 20% as the test set. The parameters *learning_rate, n_estimators, max_depth, min_child_weight, gamma, colsample_bytree, subsample* and *reg_alpha* were tuned using *RandomizedSearchCV* function. *RandomizedSearchCV* creates *n*-sets of parameters (*n=*1000 in our case) with each parameter value being selected from the list of different values that are to be considered for tuning. Models are developed using these parameters by training on the chosen data set. Each model’s predictive ability is evaluated using a 5-fold cross-validation analysis and the best set of parameters is selected based on the average of mean squared error for the validation sets. The parameters that yielded the lowest validation error for our data set were *learning_rate*=0.04, *n_estimators*=1300, *max_depth*=3, *min_child_weight*=15, *gamma=*0, *subsample*=0.8, *colsample_bytree*=0.85, *reg_alpha*=0.0001. With these parameters, the RMSEs for the training and test sets were 0.0279 and 0.0320, respectively. The quality of prediction seemed to be robust against changes in the training set as shown by the 5-fold cross-validation analysis, with a standard deviation of 0.0005 and 0.0029 for the training and the test set RMSEs.

### Individual variable contributions

Contribution of each variable towards each prediction was obtained through SHAP analysis of the model with the tuned parameters. The Python implementation of SHAP (https://github.com/slundberg/shap) was used for our interpretable AI analyses as well as for generating figures. The analyses were repeated with a 5-Fold cross-validation and the differences were found to be minimal.

## Data Availability

All analyses were based upon publicly available data.
NO PATIENT DATA was used in the analyses.
All scripts and analyses will be made available upon request.

## Author contributions

CKS, MRB performed the machine learning calculations; MRB, HJ, BS extracted the epidemiological data and performed analyses of rates; CKS, MRB, HJ, BS, MKP analyzed the results; MKP conceived the project, curated the data on government policies and wrote the paper;

## Supplementary Data Availability

The source data, and figures for all states are available at https://github.com/meherpr/COVIDrates

## Competing interests statement

The authors declare no competing interests.

## Supplementary Figures

**Supplementary Figure 1.**
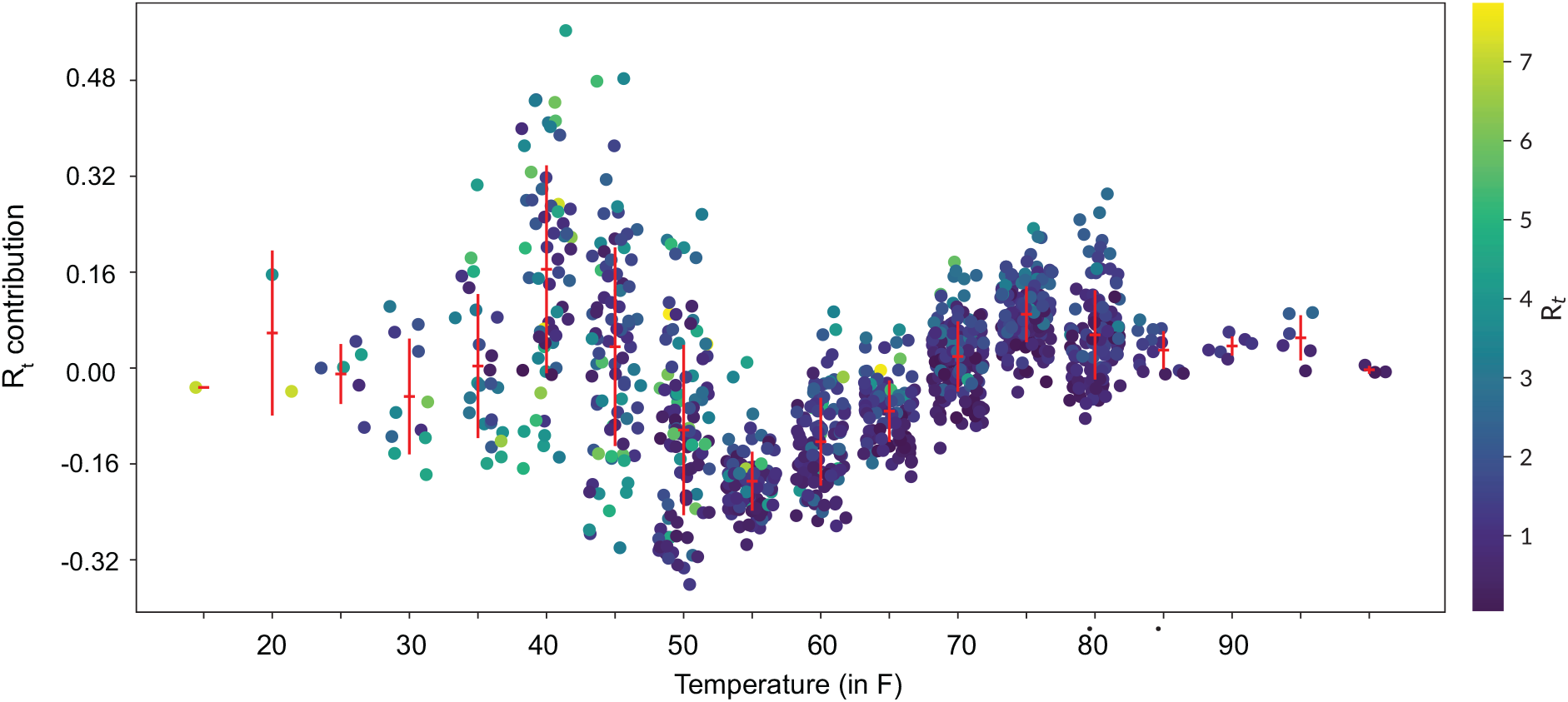
Contributions to the reproduction number using temperature as one of the variables. The effect is cannot be clearly inferred as one of reducing transmission with temperature. As discussed in the main text, this variable was subsequently discarded and was replaced by two variables HDD55, CDD55 which represent the heating and cooling needs at a given temperature.

**Supplementary Figure 2.**
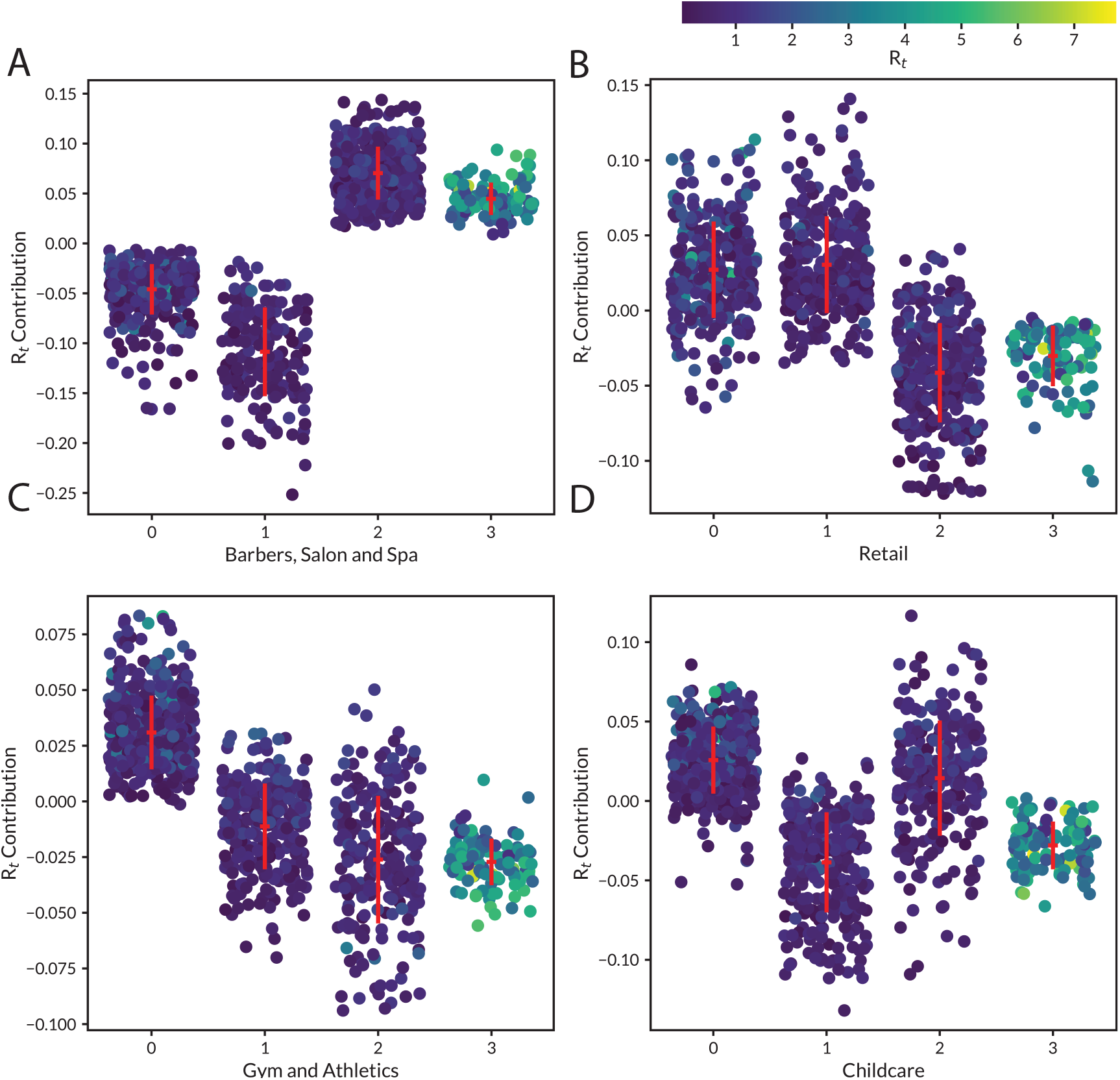
The contribution of the effects of opening various second-level of essential services is shown.

**Supplementary Figure 3.**
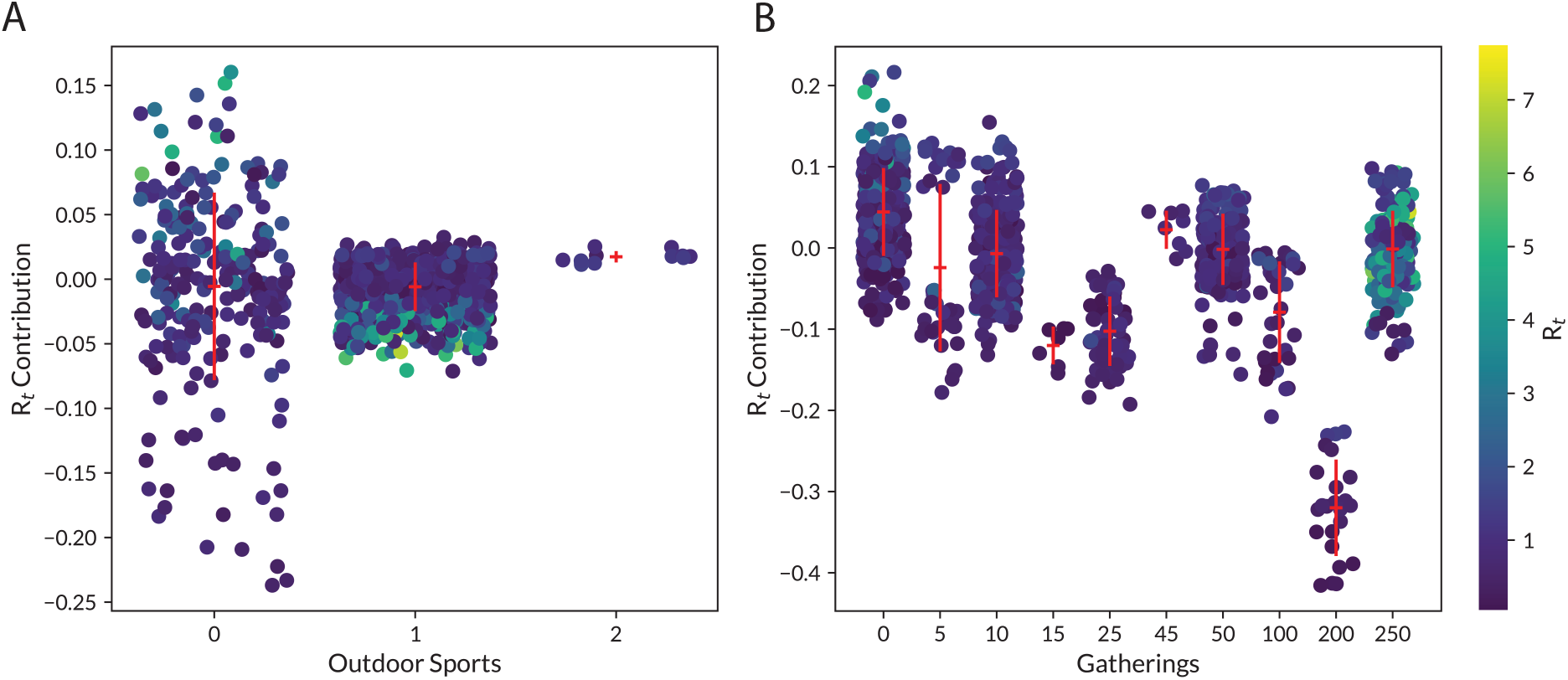
The contributions to R_t_ from outdoor events such as sports or gatherings (while respecting social distancing norms) is shown.

**Supplementary Figure 4.**
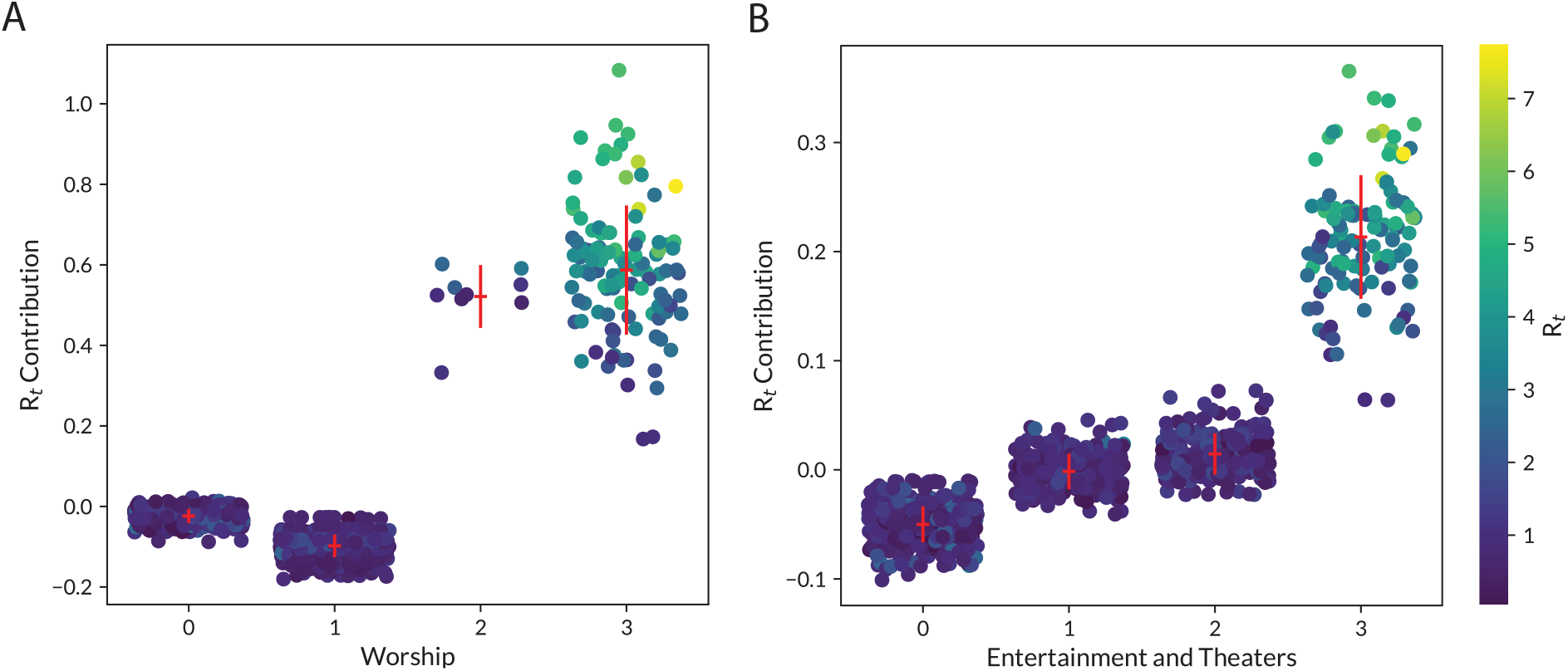
The contributions to Rt from the opening of places of worship, and entertainment theaters is shown.

## Notes

### Competing Interest Statement

The authors have declared no competing interest.

### Funding Statement

No funding was received.

### Author Declarations

No clinical data was used. Ethics approval is not applicable

